# Demonstrating the potential of untargeted hair proteomics for personalized biomarkers in stress-associated disorders

**DOI:** 10.1101/2025.03.10.25323673

**Authors:** M. Sicorello, J.-C. Sprenger, L. Störkel, B. Sarg, L. Kremser, C. Schmahl, I. Niedtfeld, A. Karabatsiakis

## Abstract

Biomarker research in psychopathology increasingly employs high-dimensional omics approaches. Yet, proteomics based on human hair remain largely unexplored, despite its potential to efficiently capture stable biological signals accumulated over weeks to months. This study leveraged machine learning to investigate the potential of the hair proteome—all detectable peptides and proteins—as a biomarker source for stress-associated psychopathology. We analyzed protein profiles from hair segments of women with non-suicidal self-injury disorder (n = 36) and healthy controls (n = 32). Of 1114 identified proteins, 611 were sufficiently abundant for analyses. Partial Least Squares Discriminant Analysis achieved stable 84.4% cross-validated accuracy for classification of clinical groups (p < .001), outperforming models based on data-derived clusters (60%), stress-related proteins (73%), and simulated hair cortisol from meta-analytic effect sizes (53–59%). Predicted class probabilities strongly correlated with clinical symptoms and well-being (r > .60). Key predictive proteins were linked to pain perception, oxidative stress, and cholesterol homeostasis. Approximately 15% of proteins differed significantly between groups, with the strongest candidates related to ribosomal function—an emerging target in depression. These findings establish hair proteomics as a promising, non-invasive biomarker source for psychiatric research, warranting validation in larger cohorts and exploration of clinical applications in risk assessment and personalized interventions.

## Introduction

About one in eight people worldwide suffer from a mental disorder (World Health Organization, 2022). From a physicalist perspective, psychopathological symptoms are shaped by underlying biological systems and their interactions with environmental influences (Insel & Cuthbert, 2015). Identifying these biological mechanisms is crucial for explaining, preventing, predicting, and treating mental disorders in a personalized manner (Cuthbert, 2014; Kapur et al., 2012). In pursuit of this goal, extensive efforts have focused on identiying biomarkers that track dysfunctions in these systems (Abi-Dargham et al., 2023; García-Gutiérrez et al., 2020).

Increasingly, biomarker research highlights the advantages of *Omics* approaches, which integrate a large number of biological features within a given modality. Combined with machine learning techniques, these approaches enhance predictive accuracy (Searles Quick et al., 2021; Sethi & Brietzke, 2016; Stolfi et al., 2024). For example, blood-based compounds have been widely studied in mental health research from an Omics perspective (Guest et al., 2016). However, biomarkers derived from body fluids are subject to rapid physiological fluctuations. A prominent example is cortisol, which follows circadian rhythms that complicate its interpretation in relation to stable mental conditions (Gotlieb et al., 2018). The high frequency of such biological fluctuations, along with other factors affecting temporal stability, presents challenges for identifying robust biomarkers of psychopathology.

Human hair represents a particularly promising source for biomarkers of psychopathology. Proteins are deposited in the hair shaft from capillaries and sweat glands via the hair follicle (Gow et al., 2010; Jang et al., 2019). As a result, hair-based biomarkers offer a retrospective, time-resolved record of physiological events that occurred during the hair growth period, with each centimeter of hair reflecting approximately one month of biological history (Kirschbaum et al., 2009; Loussouarn et al., 2016). This time-resolved property is especially advantageous in clinical settings, for example, when investigating biological changes that precede symptom onset or when biomarker concentrations in blood or saliva are too susceptible to short-term influences. Moreover, hair samples have the additional advantage of being easy to collect, transport, and store (Jang et al., 2019). This allows their easy implementation in clinical care for diagnosis, prognosis, and longitudinal observation.

Despite its potential, *Omics*-based approaches to hair biomarkers have only recently begun to emerge, and have mainly been applied in fields such as forensics, archaeology, and medical hair condition, mostly focusing on individual compounds rather than machine learning methodology (for a review see Jang et al., 2019). In psychopathology research, a hair proteomics approach is entirely lacking. Here, hair-based biomarkers have focused on single biological compounds, with the most extensive work examining hair cortisol as an index of hypothalamic-pituitary-adrenal (HPA) axis activity (Kirschbaum et al., 2009). Although the HPA axis is a major target in stress-associated conditions characterized by emotion dysregulation—such as depression, non-suicidal self-injury disorder (NSSID), borderline personality disorder (BPD), and posttraumatic stress disorder (PTSD) — meta-analyses have found that associations between hair cortisol and these disorders are mostly small or non-existent, demonstrating the limits of this biomarker (Kaess et al., 2021; Khoury et al., 2019; Psarraki et al., 2021; Schumacher et al., 2019; Thomas et al., 2019). This raises the question whether and how hair-based biomarkers can meaningfully contribute to the explanation and prediction of psychopathology.

To address this, we present a machine learning-based proteomics approach to evaluate the potential of the hair proteome as a source for biological signatures related to stress-associated psychopathology. While there are many viable stress-associated disorders for this goal, we chose NSSID as a heterogeneous clinical group sharing the feature of unspecific intense emotional distress, which most commonly motivates the self-injurious acts and urges (Hepp et al., 2020). Using this clinical group, we investigated the following key questions:

1. **Prediction (Q1):** Can a machine learning model based on the hair proteome distinguish individuals with NSSID from controls without mental disorders?
2. **Interpretation (Q2):** Can this model be interpreted in terms of (a) psychological symptoms and (b) molecular functions?
3. **Dependency (Q3):** What is the minimum number of proteins necessary and sufficient for optimal predictive accuracy?
4. **Superiority (Q4):** Does this model outperform competing models based on (a) stress-related proteins alone, (b) cortisol levels derived from simulations on meta-analytic effect sizes, (c) empirically derived protein clusters, and (d) hair-wide bivariate associations with single proteins?

By addressing these questions, the present study aims to gain first insights into the feasibility of the human hair proteome as a biomarker for mental disorders, stimulating and informing a novel research program.

## Methods

### Participants and Procedure

Participants were 68 women (n=36 NSSID, n=32 women without current mental health diagnosis [“healthy controls”; HC]) recruited using flyers, local advertisements, advertisements in social media and the website of the institute, NSSID-related Facebook groups, an existing participant pool, and direct contact with patients on the waitlist for treatment at the Central Institute of Mental Health (Mannheim, Germany). Inclusion criteria for the NSSID group comprised women aged 18–45 years who met DSM-5 criteria for NSSID, operationalized as engaging in at least one self-injury per week over the preceding three months. General exclusion criteria included a body mass index (BMI) < 17.5 or > 34.0 kg/m^2, a lifetime diagnosis of schizophrenia, intellectual disability, cognitive impairment, substance dependence within the past six months, pregnancy, or current use of opiates, corticosteroids, or naltrexone. Mental health status in both groups was assessed using the Structured Clinical Interview for DSM-5 (SCID-I) and the German version of the International Personality Disorder Examination (IPDE; Loranger et al., 1997). All participants completed the Borderline Symptom List (BSL; Bohus et al., 2009) and the German version of the Childhood Trauma Questionnaire (CTQ; Wingenfeld et al., 2010). For descriptive statistics see Table 1. All participants provided written informed consent and received financial compensation. This study was approved by the ethics committee of the Medical Faculty Mannheim, Heidelberg University (2014-601N-MA)

**Table 1.** Descriptive sample characteristics.

| | NSSID<br>($M \pm SD$) | Control<br>Group<br>($M \pm SD$) | $p$-value | Cohen's $d$ |
| --- | --- | --- | --- | --- |
| Age | 23.56 ±<br>6.28 | 28.42 ±<br>6.47 | 0.003 | -0.76 |
| BMI | 24.74 ±<br>4.73 | 22.45 ±<br>3.36 | 0.024 | 0.55 |
| CTQ Total score | 58.72 ±<br>20.08 | 29.69 ±<br>7.15 | 0.000 | 1.88 |
| BSL: BPD symptoms | 58.72 ±<br>15.99 | 1.96 ±<br>2.78 | 0.000 | 4.61 |
| BSL: Dysfunctional<br>behavior | 5.67 ±<br>4.06 | 0.31 ±<br>0.55 | 0.000 | 1.72 |
| BSL: Current well-being | 3.03 ±<br>1.65 | 8.27 ±<br>1.40 | 0.000 | -3.38 |
| BPD | 24 |  |  |  |
| Major Depression | 23 |  |  |  |
| PTSD | 15 |  |  |  |
| Social Phobia | 7 |  |  |  |
| Specific Phobia | 4 |  |  |  |
| Anorexia Nervosa | 4 |  |  |  |
| Bulimia Nervosa | 4 |  |  |  |
| Other disorders | 12 |  |  |  |
| SS(N)RI | 15 |  |  |  |
| Other Antidepressants | 5 |  |  |  |
| Neuroleptics | 12 |  |  |  |

*Note.* BMI = Body Mass Index, CTQ = Childhood Trauma Questionnaire, BSL = Borderline Symptom List, BPD = Borderline Personality Disorder, PTSD = Posttraumatic Stress Disorder, SS(N)RI = Selective Serotonin (Noradrenalin) Reuptake Inhibitor.

### Hair Sample Collection and Preparation

Hair samples were collected from the posterior vertex position of the scalp as reported previously (Karabatsiakis et al., 2022) following a standard procedure. A hair strand approximately 5–7 mm in diameter was secured with thread and cut as close to the scalp as possible using sterilized scissors and clean gloves. Samples were wrapped in aluminum foil, labeled, and stored at -80°C at the BioPsy Biobank of the Department of Genetic Epidemiology at the CIMH Mannheim (Witt et al., 2016) until preanalytical procedures. After completing the hair sampling, all hair strands were segmented into proximal (1 cm, representing the most recent month; Kintz et al., 2006) and distal (1 cm, representing the preceding month) sections. Segments were weighed, pulverized using a Retsch Mixer Mill MM 301 (3 x 5 min, 30 Hz) with liquid nitrogen cooling cycles between the runs, and centrifuged to concentrate the powdered hair. Approximately 0.72 ± 0.18 mg of hair powder per sample was allocated for protein extraction, following the protocol of (Carlson et al., 2018).

### Hair Sample Collection and Preparation

Hair samples were collected from the posterior vertex position of the scalp as reported previously (Karabatsiakis et al., 2022) following a standard procedure. A hair strand approximately 5–7 mm in diameter was secured with thread and cut as close to the scalp as possible using sterilized scissors and clean gloves. Samples were wrapped in aluminum foil, labeled, and stored at -80°C at the BioPsy Biobank of the Department of Genetic Epidemiology at the CIMH Mannheim (Witt et al., 2016) until preanalytical procedures. After completing the hair sampling, all hair strands were segmented into proximal (1 cm, representing the most recent month; Kintz et al., 2006) and distal (1 cm, representing the preceding month) sections. Segments were weighed, pulverized using a Retsch Mixer Mill MM 301 (3 x 5 min, 30 Hz) with liquid nitrogen cooling cycles between the runs, and centrifuged to concentrate the powdered hair. Approximately 0.72 ± 0.18 mg of hair powder per sample was allocated for protein extraction, following the protocol of (Carlson et al., 2018).

### Preanalytical sample preparation for mass spectrometry proteomics analysis

Exact amounts of hair (ranging from 0.5 to 2.5 mg) were suspended in 120 µL extraction buffer – consisting of 100 mM ammonium bicarbonate (ABC) buffer pH 8, 8 M urea, and 50 mM dithiotreitol (DTT) – and shook at room temperature for 24 hours. The proteins were reduced by adding 12 µL of 100 mM DTT (1 h at RT), alkylated with 120 µL 50 mM IAA (1 h at RT), and quenched with 120 µL 100 mM DTT. Samples were diluted by adding 828 µL ABC buffer and 120 µL acetonitrile to reduce the concentration of urea below 1 M. An amount of 0,8 µg Trypsin (Promega) per 1 mg hair powder was added to the samples and incubated at 37 °C for 18 h. The volume of the digested samples were reduced on a SpeedVac to 100 µL, centrifuged at 36,000 g for 10 min, and the supernatant purified by ZipTip C18 pipette tips (Thermo Scientific), according to the manufacturer’s instructions. An aliquot corresponding to 20 µg hair of digested samples was subjected to nanoLC-MS analysis.

Samples were analyzed using an UltiMate 3000 nano-HPLC system coupled to an Orbitrap Eclipse mass spectrometer (Thermo Scientific, Germany). The peptides were separated on a homemade fritless fused-silica microcapillary column (100 µm i.d. x 280 µm o.d. x 16 cm length) packed with 2.4 µm reversed-phase C18 material (Reprosil). Solvents for HPLC were 0.1% formic acid (solvent A) and 0.1% formic acid in 85% acetonitrile (solvent B). The gradient profile was as follows: 0-4 min, 4% B; 4-57 min, 4-35% B; 57-62 min, 35-100% B, and 62-67 min, 100 % B. The flow rate was 300 nL/min.

The Orbitrap Eclipse mass spectrometer equipped with a field asymmetric ion mobility spectrometer (FAIMS) interface was operating in the data dependent mode with compensation voltages (CV) of -45, and -65 and a cycle time of one second. Survey full scan MS spectra were acquired from 375 to 1500 m/z at a resolution of 120,000 with an isolation window of 1.2 mass-to-charge ratio (m/z), a maximum injection time (IT) of 50 ms, and automatic gain control (AGC) target 400 000. The MS2 spectra were measured in the Orbitrap analyzer at a resolution of 15,000 with a maximum IT of 22 ms, and AGC target or 50 000. The selected isotope patterns were fragmented by higher-energy collisional dissociation with normalized collision energy of 30%.

Data Analysis was performed using Proteome Discoverer 2.5 (Thermo Scientific) with search engine Sequest. The raw files were searched against the UniProt human database. Precursor and fragment mass tolerance was set to 10 ppm and 0.02 Da, respectively, and up to two missed cleavages were allowed. Carbamidomethylation of cysteine was set as static modification, oxidation of methionine was set as variable modifications. Peptide identifications were filtered at 1% false discovery rate.

The molecular functions of proteins provided by Proteome Discoverer 2.5 were complemented with a manual search on uniprot.org to provide missing primary and secondary functions.

### Statistical Analysis

#### Preprocessing

Only compounds with missing values in less than 50% of participants in both groups were used for statistical analyses. Median imputation was used on missing values in the remaining compounds. The influence of median imputation on model accuracy was tested in sensitivity analyses. All compounds were subjected to a box-cox transformation as an objective approach to reduce skewness in the distributions and then z-transformed.

#### Machine Learning Procedure

*Partial Least Squares Discriminant Analysis* (PLS-DA) from the *caret* package (Kuhn, 2008) in R (v4.3.2) was applied as the main algorithm, which reduces the number of predictors to a smaller set of components by also taking into account information on the group criterion variable. This algorithm is particularly well-suited for high-dimensional settings with high intercorrelations between predictors and commonly used for biological data (Müller et al., 2023). The model was built using a 5×5 nested cross-validation procedure. The five inner cross-validation folds were used to determine the optimal number of components with a grid search from one to 30 components. The five outer cross-validation folds were used to determine the model accuracy based on this optimal number of components. The outer folds were stratified for group membership to ensure enough observations of both groups are present. This procedure was repeated five times for stability. The average percentage of correct classifications across all five repetitions was calculated as the main accuracy metric.

The same PLS-DA model was used to determine the number of necessary and sufficient compounds for prediction, the only difference being that the inner cross-validation folds were used to pick the best predictors instead of the optimal number of components (which was fixed to the number of components in the full model).

#### Cluster Analysis

Data-driven clusters were identified to inform the structure of the hair proteome and check whether focusing on a small number of clusters is superior to using the unrestricted hair proteome. Two complementary clustering approaches were used. First, hierarchical agglomerative clustering was performed on a matrix of Euclidean distances using Ward’s method. The number of clusters was identified based on the elbow method for within-cluster sums of squares and the silhouette method using the package factoextra. Second, Density-Based Spatial Clustering of Applications with Noise was used with the R package *dbscan* (Hahsler et al., 2019). While the hierarchical agglomerative clustering approach clusters all data points until all are in the same cluster, dbscan only clusters data points based on two prespecified hyperparameters (minPts, the minimum cluster size, and ε, the radius around a given data point that is searched for other points). This allows distinguishing between clusters of proteins and single proteins that do not belong to a coherent cluster, which are summarized into a “noise” category. The hyperparameter ε was chosen by selecting different values for minPts (3, 5, and 8) and identifying the knee in each plot of k-nearest-neighbor distances between all points. The y-axis value at the knee is the chosen hyperparameter ε.

### Open Science Practices

Analysis code and modelling outputs can be found on github: https://github.com/MaurizioSicorello/NSSI_hairProteomics. As the identifiability of participants based on the quantitative hair proteome is unclear but not unlikely, data access can only be provided based on individual requests in compliance with the European General Data Protection Regulation. A table of identified proteins and their molecular functions can be found in the github repository (“results/PLS_varImpAndFunctions.csv”).

## Results

We identified 611 proteins which were available in at least 50% of samples in each of the two groups. A subset of 201 proteins could be identified without missing data on any participant, which is a desirable property for clinical application.

### Prediction (Q1): Proteome-wide predictive models

#### Model building and evaluation

The PLS-DA model exhibited relatively good and statistically significant classification accuracy (accuracy = 84.4%, *p* < .001). Most models in the repeated nested cross-validation procedure favored a solution with three latent components (12 out of 25 models), followed by four latent components (9 out of 25). The three-component model showed a good balance of cross-validated sensitivity (84.3%) and specificity (81.1%). The receiver operating characteristic curve indicated that even at a classification threshold with an estimated specificity of 100%, over 60% of NSSID cases are still detected (Figure S1). This is a relevant diagnostic scenario for large-scale screenings in populations predominantly without mental disorders. The predictive accuracy was maintained when predictors with missing values were removed instead of imputed according to different missingness thresholds (Figure S2).

A random forest algorithm, which in contrast to PLS-DA allows for nonlinearities and interaction effects, did not improve upon classification, with lower accuracies for all evaluated hyperparameters (accuracies 70-78% for feature selection hyperparameters [*mtry*] 1-200). Therefore, we will focus on the PLS-DA model with three components in the following analyses.

### Interpretation (Q2)

#### Psychological interpretation

We explored which dimensional features were most strongly reflected in the full PLS-DA model by correlating (a) the predicted probability to belong to the NSSID class and (b) the participant scores on the three PLS components with (1) childhood maltreatment, (2) BMI, (3) age, and (4) BPD psychopathology. For borderline psychopathology, we distinguished between the three BSL scales for symptoms, dysfunctional behavior, and overall well-being during the last week. Importantly, class probabilities for each participant were calculated from leave-one-out cross-validation to prevent inflated correlations due to overfitting.

The model’s predicted class probabilities were most strongly correlated with BPD psychopathology (Figure 1). The correlations with all three BSL subscales exceeded *r* = .50, with particularly strong correlations for symptoms and general well-being during the last week (*r* > .60). The findings for general well-being are especially meaningful, as this scale had approximately equal variance in both groups, while the other two BPD scales show clear floor effects in the HC group (Table 1). Class probabilities were also highly correlated with childhood trauma and age, albeit to a lesser degree (|*r|* > .40).

**Figure 1.**
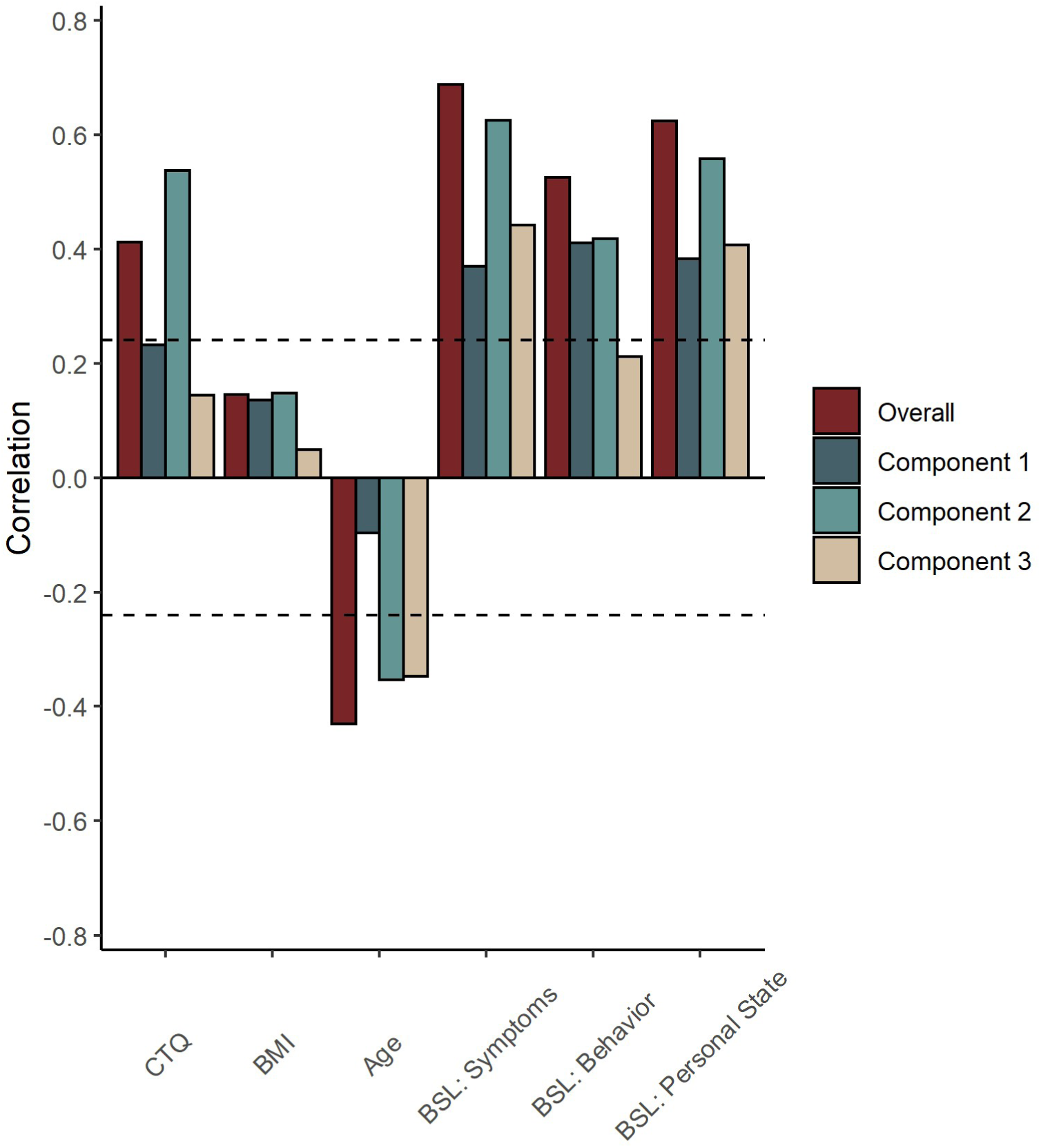
Correlations between leave-one-out predictions (class probabilities) and other covariates. Class probabilities are shown based on predictions by the full model and separately for each of the three model components. CTQ = Childhood Trauma Questionnaire, BDI = Body Mass Index, BSL = Borderline Symptom List.

To ensure the model’s good accuracy is not due to confounding by group differences in age (Table 1), we matched the mean age of the two samples more closely and re-ran the full PLS-DA model. First, we removed the five youngest NSSID participants to reach equal group sizes. Then, we iteratively removed the youngest NSSID and the oldest HC participant until we reached a mean age gap below 1.5 years. This led to a sample of 54 participants with a group difference in age of 1.4 years. In this age-matched sub-sample, the predictive accuracy of the full PLS-DA model was still maintained (81.5%). Interestingly, despite matching for age, the predicted class probabilities were still substantially correlated with age: *r*(53) = -.30, *p* = .029. Hence, the PLS-DA model is likely not compromised by confounding with age, despite age being substantially associated with the model predictions.

#### Molecular interpretation

In total, we assigned 68 different primary molecular functions to the 611 proteins. Table 2 (column 1) shows the primary functions of the top ten most important predictors. “Translation” and “structural protein (hair/nails/epidermis)” were five of the top ten contributing predictors. Still, these were also the most frequently assigned primary functions overall (21% of proteins), which might lead to an overestimation of functional importance by chance. Therefore, we calculated the average variable importance per primary function, which controls for their base rates (Table 2, column 2). Here, translational and structural functions were no longer represented. Instead, some noteworthy molecular functions included pain perception (Protein NipSnap homolog 1), implicated in NSSID, as well as cholesterol homeostasis and oxidative stress response, implicated in psychopathology more generally.

**Table 2.**
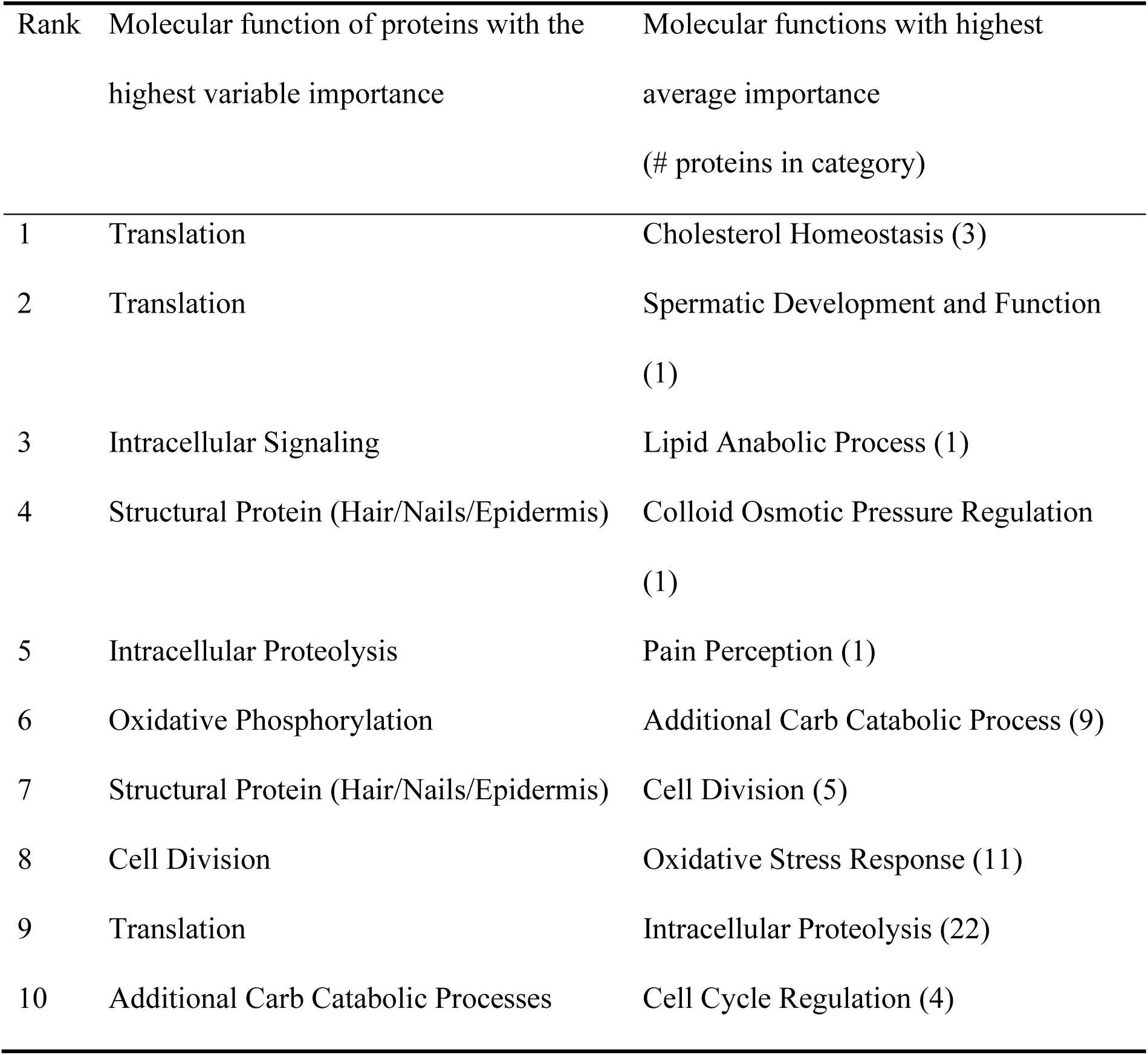
Most important protein functions in the predictive model.

Importantly, while the table only shows primary functions, single proteins mostly have multiple secondary functions. For example, the second most predictive molecular function was “Spermatic Development and Function”, which seems counterintuitive in a female sample. Nevertheless, this was due to only a single protein (*Tetratricopeptide repeat protein 21A*) which is also involved in protein transportation within cilia. A full table of sorted variable importances together with functional characteristics (e.g., primary and secondary functions) can be found in the github repository (see section on Open Science Practices).

### Dependency (Q3): Number of necessary and sufficient metabolites

While the full PLS-DA model with all 611 metabolites had good predictive accuracy, the number of necessary proteins might be much lower. This was already partially indicated by our analysis on the impact of missing data imputation (Figure S2).

We explored this possibility more systematically by testing how many of the most predictive candidates (a) were *sufficient* to reach a plateau in cross-validated accuracy and (b) can be removed while predictive accuracy is still maintained (i.e., are *necessary* for prediction). As can be seen from the blue line in Figure 2, already the 10 most predictive proteins were sufficient to reach an independently cross-validated accuracy over 80%. There was only a negligible increase around 2% when approximately 250 proteins were included, followed by a plateau. Still, while only a few proteins appeared to be sufficient for accurate prediction, they are not necessary: Even when the best 100 proteins were removed, the model maintained its full cross-validated accuracy (red line). A steep decline was only observed from the removal of 200 proteins onwards. Randomly choosing proteins only reaches similar accuracies when at least 25% of proteins are included (black dashed line). Hence, there is useful information for classification distributed across most but not all parts of the hair proteome, which can already be sufficiently contained in a small set of proteins.

**Figure 2.**
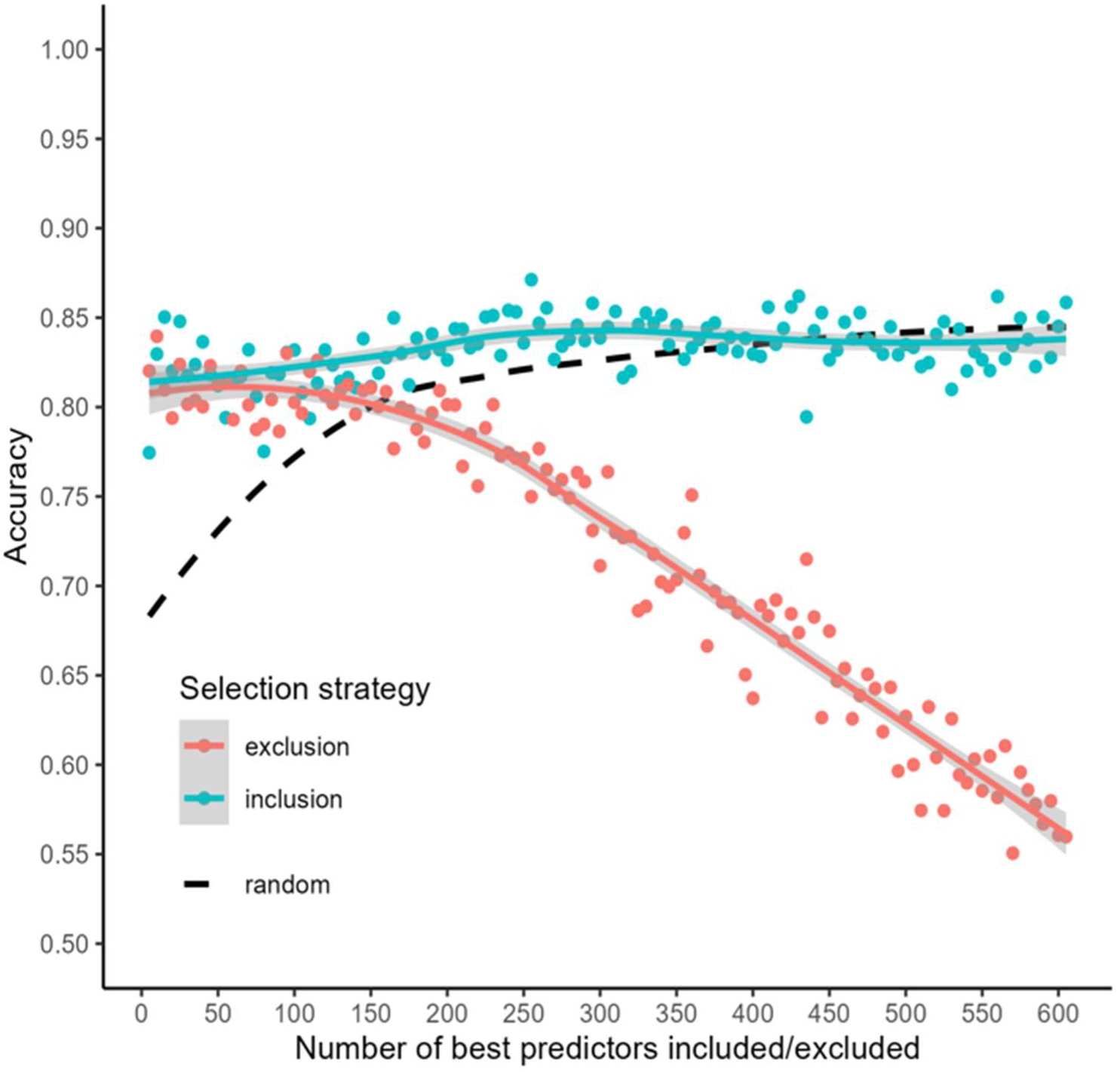
Cross-validated accuracies by different predictor selection strategies. For the inclusion strategy, the x best predictors were chosen based on training folds and a retrained model only including these predictors evaluated in the test fold. For the exclusion strategy, the x best predictors were excluded. The black dashed line represents accuracy for random draws of x predictors as a reference, averaged over 25 repetitions for stability.

### Superiority (Q4): Alternative Predictive Models

#### Performance of stress-related proteins

We identified 112 proteins linked to the generalized molecular stress response via primary or secondary functions (e.g., Alpha-crystallin B chain, Heat shock protein 27, Stomatin-like protein 2). A PLS-DA model including only these stress-related proteins had substantially lower accuracy (72.9%). In contrast, when these proteins were removed from the full model, the original accuracy was maintained (82.6%). Hence, proteins known to be involved in the stress response are neither necessary nor sufficient for good prediction.

For comparison with hair cortisol, the most frequently investigated hair metabolite in psychopathology research, we simulated what the expected cross-validated accuracy would have been, if we had used only hair cortisol with logistic regression to predict group membership. We simulated 1000 iterations of normally distributed data with our study’s sample size and a population group difference of *d* = .213 (overall) and *d* = .478 (optimistic) based on a recent meta-analysis on the association between adversity and hair cortisol (Khoury et al., 2019). The two effect sizes corresponded to an average accuracy of only 53.3% (*SD* = 5.8) and 58.7% (*SD* = 6.4). This would be practically identical for a larger sample of *N* = 500 and equal group sizes, with accuracies of 54.1% (*SD* = 2.4) and 59.4% (*SD* = 2.1).

In sum, focusing on cortisol in hair and many other proteins related to generalized molecular stress might not be sufficient to reach acceptable predictive accuracies for psychopathology.

#### Empirical protein clusters

Hair protein levels were overall positively correlated to a considerable degree, indicating that they might be structured in covarying clusters (*M_r_* = .38, *SD_r_* = .24; Figure S3). The silhouette and elbow method favored a two-cluster solution for the hierarchical agglomerative clustering approach (figure S4). Still, within-cluster correlations were relatively modest (Cluster 1: *M_r_* = .49, *SD_r_* = .21; Cluster 2: *M_r_* = .33, *SD_r_* = .21). Similarly, the *dbscan* algorithm indicated the existence of only one large cluster comprising almost all metabolites (589-593), classifying the remaining few metabolites as outliers without cluster membership.

A naïve baseline model only using average protein levels in the two clusters from the hierarchical agglomerative clustering approach did not significantly predict group membership using cross-validated logistic regression (Accuracy = 60.4%, *p* = .537).

#### Single proteins

Complementary to the machine learning-based predictive approach, we performed hair proteome-wide analyses to test for single metabolites with significant group differences using independent *t*-tests. Such metabolites can be used to develop novel hypotheses targeting biological systems. In total, 101 hair proteins showed significant group differences after FDR correction (*q* = .05), spanning 37 primary molecular functions. Statistics and molecular functions for all single proteins can be found in the online repository (“results/tableS1_bivariateAssociations.csv”). Figure 3 shows the distribution of Cohen’s *d* together with the five-fold cross-validated predictive accuracies for the single proteins from logistic regression to allow comparisons with the PLS-DA approach. Most statistically significant proteins (82%) had a cross-validated accuracy below 70%. Qualitatively, we noticed that the three proteins with the largest effect sizes are all functionally involved in ribosomal biosynthesis and had smaller concentrations in the NSSID sample (large ribosomal subunit protein uL23 at 88% accuracy, small ribosomal subunit protein eS25 at 81% accuracy, small ribosomal subunit protein uS11 at 79% accuracy). This matches recent findings that glucocorticoid responses to stress can down-regulate the expression of ribosomal protein genes in post-mortem tissue of depressed individuals and stress-exposed mice, which was reversed during remission (Zhang et al., 2023).

**Figure 3.**
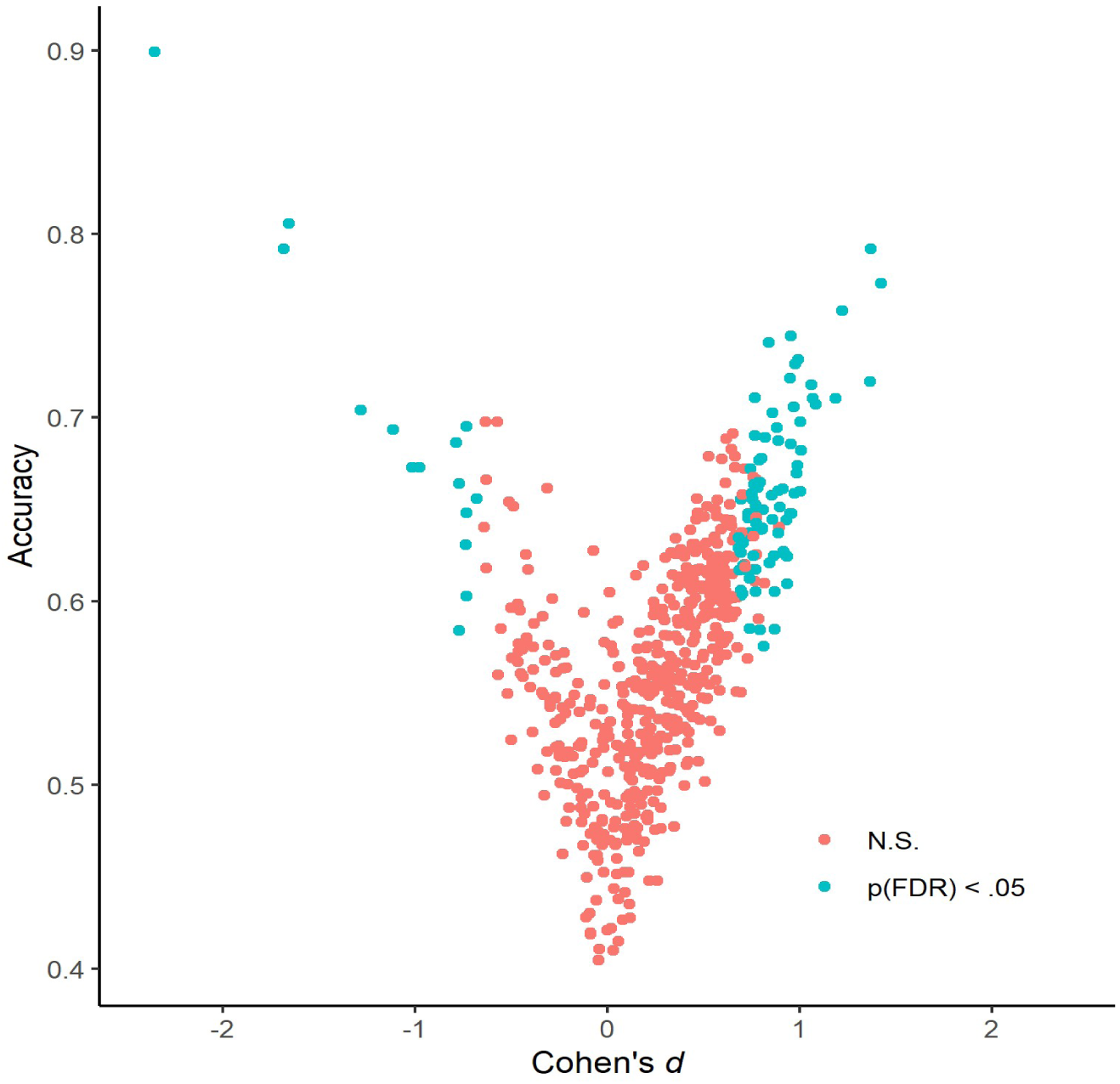
Cohen’s *d* and cross-validated prediction accuracy for single proteins. Colours indicate statistical significance after FDR correction.

Notably, very strong effect sizes for single proteins might have capitalized on chance due to the larger number of tested proteins. Hence, smaller effect sizes would be expected in future studies. Moreover, the continuous distribution of effect sizes seen in Figure 3 could suggest that many more proteins might show significant differences in designs with higher statistical power.

## Discussion

In the present study, we showcase the potential and viability of hair proteomics for biomarker identification in (stress-associated) psychopathology. In our example use-case of classifying women with NSSID versus healthy controls, the hair proteome-based machine learning models showed good predictive performance (Q1), which was stable across a range of sensitivity analyses (missing value imputation, different feature sets, subsampling for age matching). This is in stark contrast to the very small accuracies which can be expected for hair cortisol, the most used hair-based biomarker in psychopathology research, based on meta-analytic effect sizes across stress-associated disorders (Kaess et al., 2021; Khoury et al., 2019; Psarraki et al., 2021; Schumacher et al., 2019; Thomas et al., 2019). Similarly, stress-related proteins were neither necessary nor sufficient for good prediction, showing the value of broadening the range of target compounds beyond theory-driven candidates.

Besides the model’s good predictive performance, we found evidence for its clinical interpretability (Q2). From a dimensional psychological perspective, the model appeared to mainly track BPD symptoms and well-being in the last week, rather than childhood maltreatment or confounds like age and BMI. From a molecular perspective, the model emphasized the importance of biological compounds involved in stress-associated psychopathology, such as cholesterol homeostasis and oxidative stress (Black et al., 2015; Gliozzi et al., 2021). Of particular interest is the predictive importance of a protein involved in the sensory perception of pain, Protein NipSnap homolog 1, potentially relating to the common alterations of pain perception in NSSID (Koenig et al., 2016). Together with the finding that roughly 15% of proteins had statistically significant bivariate differences between groups after multiple comparison correction, this demonstrates that the hair proteome represents a rich source not only for prediction, but also for stimulating research following hair proteins towards up-stream dysregulated biological systems. This might provide accessible health-monitoring capabilities, especially for those with limited access to local health infrastructure necessary to collect blood or other biofluids.

A major follow-up question was how many proteins are necessary and sufficient for a good predictive model (Q3). This is especially relevant as studies might differ in their detected array of biological compounds. Here, we found that a small number of ten proteins can already be sufficient to reach a cross-validated accuracy plateau in the current sample. Strikingly, even when removing the first 200 most predictive features, the accuracy remained basically unchanged. This is likely due to large redundancies between proteins, which are advantageous for component-based algorithms like PLS and could be responsible for the stability we observed in our sensitivity analyses. Hence, it is not the case that a small exclusive set of biological compounds is responsible for good prediction and, in turn, using a large array of proteins might reduce noise and lead to more stable prediction.

There are two possible explanations for the observation of predictive redundancy. Firstly, disorder-associated processes (e.g. emotion dysregulation) might disrupt a large variety of biological processes, for example, due to stress, disorder-specific factors, or other clinical, demographic, and lifestyle confounds. Secondly, such factors might influence the degree to which biological compounds are deposited in the hair more generally. Importantly, these two explanations are not mutually exclusive, but together likely best account for the observations made here: On the one hand, there appears to be considerable redundancy between a large set of proteins. On the other hand, significant group differences in protein levels were found for both directions (Figure 3) and focusing only on data-driven protein clusters or 200 stress-associated proteins leads to much lower accuracies (Q4). Therefore, it will be crucial to distinguish which protein aberrations are due to specific factors versus a general compound deposition tendency where studies focus on molecular interpretation (i.e., inference regarding specific dysregulated biological systems).

Our study was intended to test the principal viability of the hair proteome as a biomarker signature for stress-associated mental disorders. Therefore, it is limited by focusing on relatively young women with NSSID as a specific inclusion criterion, which, like most disorders, has a large range of comorbidities. Due to the known limitations of most categorical taxonomies of psychopathology, especially in biomarker research, we do not believe that the main long-term goal should be to develop hair-based classifiers of specific stress-associated mental disorders (Clark et al., 2017; Insel & Cuthbert, 2015; Kotov et al., 2017). Rather, the largest applied potential might lie in a precision medicine approach towards (a) prospective “black box” prediction (e.g., of risk and symptom trajectories), (b) objective retrospective analysis of symptom-related physiology in the period before anamnesis, and (c) following concrete observable psycho-behavioral processes and symptoms back to the up-stream dysfunction of specific biological systems. Hair-based indicators of such systems might be used for personalized risk assessment and the development of biology-based treatments for mental health conditions (Comes et al., 2018). Such a research agenda should focus on identifying confounders that might drive associations between the hair proteome and psychopathology, which we could only provide to a very limited degree and are often not accounted for in the literature (e.g., specific medication, lifestyle factors, ethnicity; Rippe et al., 2016).

Overall, our study demonstrates the large potential of the hair proteome for both predicting and understanding psychopathology. It offers many insights on how the hair proteome is structured and, following from this, which machine learning approaches might lead to the best results. These insights can aid an emerging literature to refine the interpretability of hair-based proteomic markers as well as adapt them to specific clinical tasks. Such tasks might include personalized assessments of risks, prospective trajectories, and medication choice.

## Data Availability

All data produced in the present study are available upon reasonable request to the authors

https://github.com/MaurizioSicorello/NSSI_hairProteomics

