## Supplemental Figures for "Demonstrating the potential of untargeted hair proteomics for personalized biomarkers in stress-associated disorders"

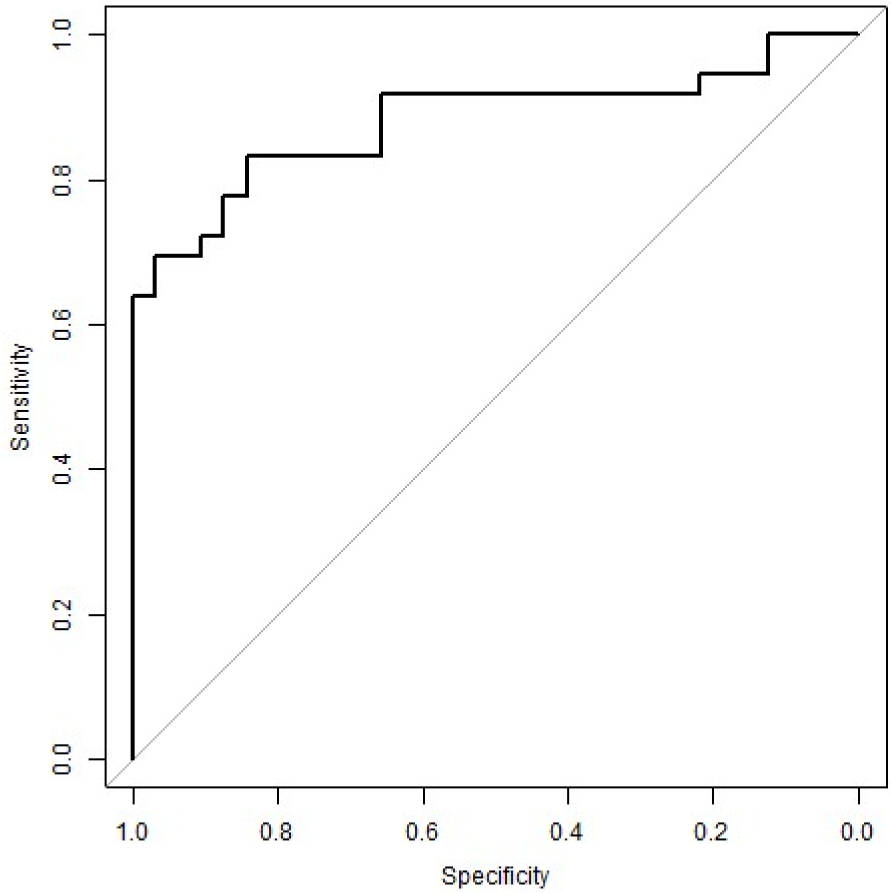


**Figure S1.** Receiver Operating Characteristic (ROC) curve of the main PLS-DA model.

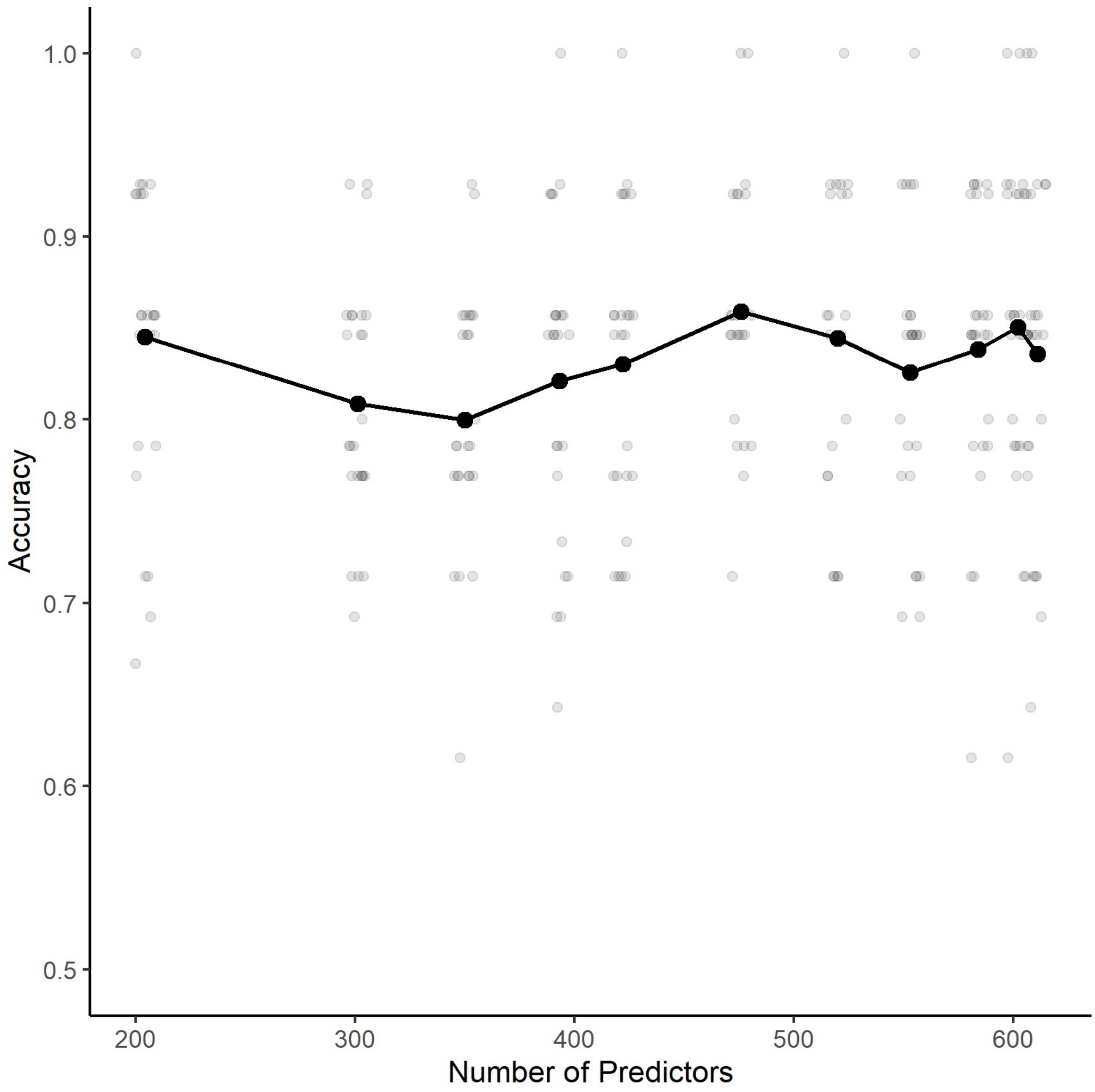


**Figure S2**. Sensitivity analysis for missing values imputation. The x-axis shows the number of predictors (i.e., biological compounds in the model), starting from using only compounds with 0% missings (200 compounds) up to allowing 50% missings (600 compounds) in 5% increments.

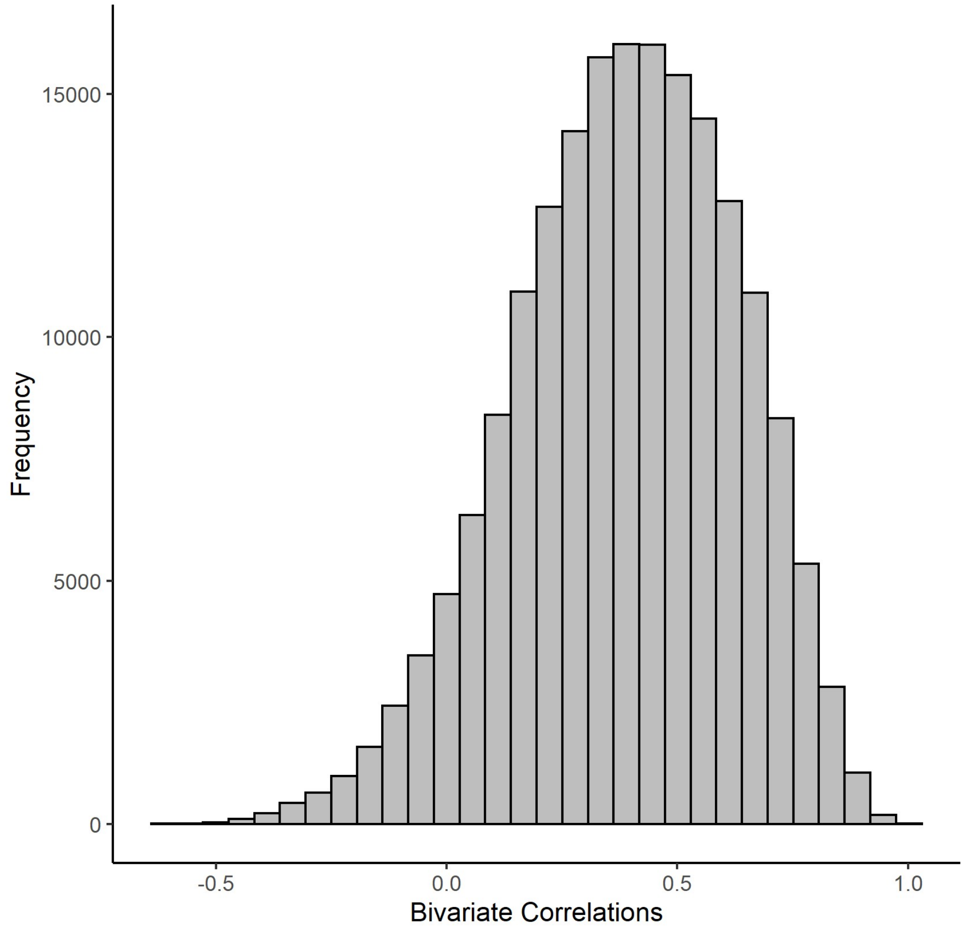


**Figure S3.** Distribution of correlations between concentrations of different biological compounds. Shows that the vast majority of compounds are positively correlated.

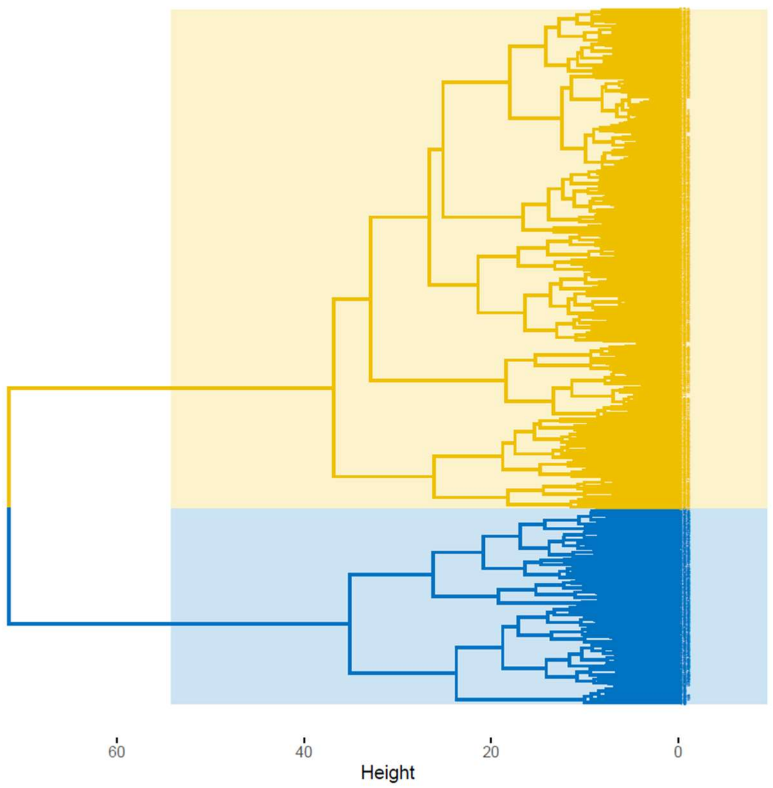


**Figure S4.** Dendrogram for the hierarchical agglomerative clustering approach.
